# EEG-based Deep Learning Reveals Cortical Sensitivity to Small Changes in Deep Brain Stimulation Parameters

**DOI:** 10.1101/2025.02.11.25321886

**Authors:** Nicolas Calvo Peiro, Mathias Ramm Haugland, Alena Kutuzova, Cosima Graef, Aminata Bocum, Yen Foung Tai, Anastasia Borovykh, Shlomi Haar

**Affiliations:** Care Research and Technology Centre, UK Dementia Research Institute, London, United Kingdom; Department of Computing, Imperial College London, London, United Kingdom; UKRI Centre for Doctoral Training in AI for Healthcare, Imperial College London, London, UK; Department of Brain Sciences, Imperial College London, London, United Kingdom; Department of Neurology, Charing Cross Hospital, London, United Kingdom; Department of Mathematics, Imperial College London, London, United Kingdom; School of Psychology, University of Surrey, Guildford, United Kingdom

## Abstract

Deep Brain Stimulation (DBS) is an invasive procedure used to alleviate motor symptoms in Parkinson’s Disease (PD) patients. Despite its effectiveness, the impact of DBS on brain activity and the way to optimise stimulation parameters remains unclear. In this study, we aimed to map the sensitivity of cortical neural response to changes in DBS parameters in real-world clinical settings. We recorded in-clinic EEG data from PD patients during their clinical DBS programming sessions, both at rest and during arm movement. A siamese variant of the EEGNet deep learning architecture was trained independently for each patient and parameter combination to determine whether two 1-sec-long EEG segments were recorded under the same or different DBS parameters, yielding 30 independently trained models. Our models achieved an average accuracy of 78% in classifying whether two 1-second EEG segments corresponded to the same or different DBS parameters. Explainability methods were then applied to extract the neural oscillations learned by the models. Ablation studies identified mid-gamma oscillations (60-90Hz) as the key band for classifying these changes. Importantly, that was not driven by 1:2□gamma entrainment. In a sub-group of patients, the theta-alpha oscillations (4-12Hz) also contributed to detecting changes in DBS parameters. We demonstrate that small DBS parameter changes modify cortical activity in a consistent manner that can be detected using shallow convolutional networks on low-density EEG. Our findings suggest that cortical mid-gamma oscillations (60-90Hz) are highly sensitive to small changes in DBS settings. Our approach and findings could be leveraged towards identifying and defining novel digital biomarkers to guide DBS programming and adaptive DBS systems, potentially improving future treatment outcomes for PD patients.

## Introduction

Deep Brain Stimulation (DBS) is an established therapy for motor symptoms in Parkinson’s Disease (PD), achieved by implanting electrodes in deep brain nuclei, typically the subthalamic nucleus (STN), and applying electrical current [1]. Programming stimulation, however, relies on a suboptimal trial-and-error process guided by clinician assessment and patient reporting [2]. The high-dimensional parameter space (amplitude, contact, frequency, pulse width) is too large to fully explore, even during extended sessions [2]. Settings remain fixed between visits, ignoring the brain’s dynamic behaviour and the patient’s fluctuating needs [3]. Adaptive closed-loop DBS (aDBS), which adjusts stimulation based on patient state, could address these limitations and reduce long-term side effects [4,5]. Early in-clinic aDBS studies [6–9] and recent in-home trials [10] show promise, but progress is constrained by the quality of digital biomarkers available to guide stimulation.

Elevated beta activity (13–30 Hz) in the STN is the most studied biomarker in PD and aDBS, [11–13], and was recently approved for clinical use following a successful clinical trial [14]. Yet, the beta peak is not evident in all patients, and is still a nosy biomarker. Recent studies have identified mid-gamma oscillations (60–90 Hz) in the STN, cortex [15], or both [10,16–19] as potential DBS biomarkers. Increased activity at half the stimulation frequency has also been observed, supporting a 1:2 entrainment mechanism for induced mid-gamma at both cortical and STN levels [16,18–21]. The entrained fine-tuned gamma (FTG) oscillations have been linked to functional improvements in symptom alleviation and finger-tapping performance [10,20]. FTG has also demonstrated translational value in an in-home aDBS trial [10], but seems to be mostly synchronised to the medication cycle.

Importantly, while existing research on biomarkers has provided insightful knowledge, only a few of those studies explored the sensitivity to small changes in the DBS parameters, which clinicians perform during programming sessions. Moreover, it was mostly conducted in settings with clinically ineffective stimulation or non-translational paradigms, leading to a lack of robustness and real-world applicability [22–24].

To address this, we took a data-driven approach using AI to find patterns in the data by looking for the sensitivity of neural oscillations to small changes in DBS parameters. We developed a Siamese Neural Network (SNN) based on EEGNet [25] to detect neural sensitivity to subtle parameter changes. The SNN was trained to classify whether pairs of 1-second EEG segments were recorded under identical or different stimulation settings, and explainability methods were applied to identify the features learned. Using data from unmodified clinical programming sessions, our approach provides real-world value. Together with the use of very short (1-sec) segments, which could enable automatic scanning of the vast DBS parameter space, it advances the search for robust digital biomarkers, towards biomarker-guided automated DBS programming [26] and aDBS.

## Methods

### Participants

We analysed a dataset collected during routine DBS programming sessions in PD patients, where clinicians adjusted parameters and assessed motor symptoms. All participants provided written informed consent, and the study was approved by the London–Harrow Research Ethics Committee in accordance with the Declaration of Helsinki. Patient demographics are shown in Table 1, and parameter ranges for each patient in Supplementary Table 1.

**Table 1.**
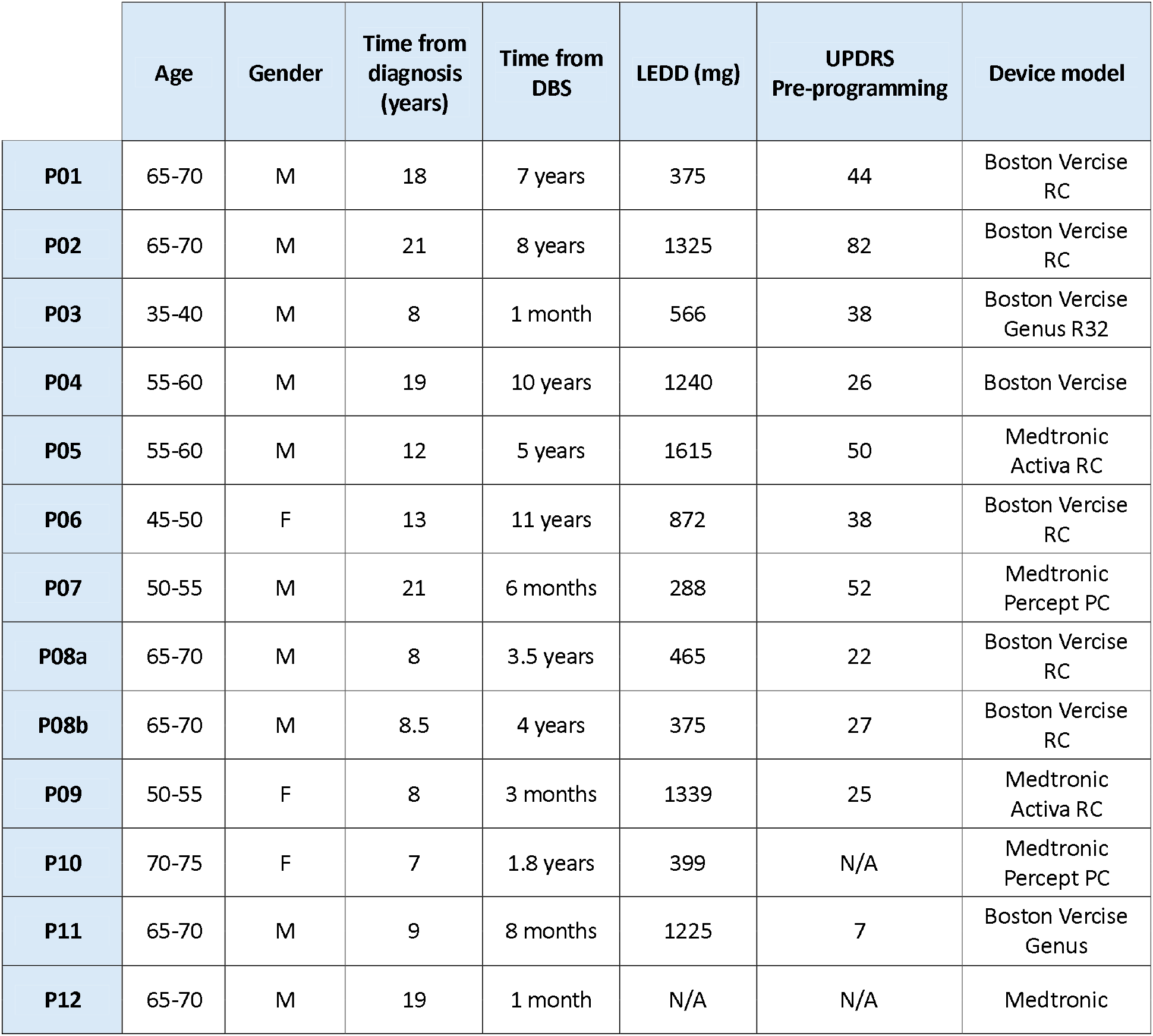
Patient characteristics. Unified Parkinson’s Disease Rating Scale (UPDRS) scores were evaluated before the start of the session. All patients, apart from P03 and P11, were recorded ON medication and while DBS was ON but not optimised. P03 and P12 were recorded during their monopolar review, indicated by the time from DBS being 1 month. The Average Tremor UPDRS score was annotated from videos of the DBS sessions and corresponds to an average of all tremor UPDRS scores given throughout the programming session. LEDD = Levodopa Equivalent Daily Dose

|  | Age | Gender | Time from diagnosis (years) | Time from DBS | LEDD (mg) | UPDRS Pre-programming | Device model |
| --- | --- | --- | --- | --- | --- | --- | --- |
| **P01** | 65-70 | M | 18 | 7 years | 375 | 44 | Boston Vercise RC |
| **P02** | 65-70 | M | 21 | 8 years | 1325 | 82 | Boston Vercise RC |
| **P03** | 35-40 | M | 8 | 1 month | 566 | 38 | Boston Vercise Genus R32 |
| **P04** | 55-60 | M | 19 | 10 years | 1240 | 26 | Boston Vercise |
| **P05** | 55-60 | M | 12 | 5 years | 1615 | 50 | Medtronic Activa RC |
| **P06** | 45-50 | F | 13 | 11 years | 872 | 38 | Boston Vercise RC |
| **P07** | 50-55 | M | 21 | 6 months | 288 | 52 | Medtronic Percept PC |
| **P08a** | 65-70 | M | 8 | 3.5 years | 465 | 22 | Boston Vercise RC |
| **P08b** | 65-70 | M | 8.5 | 4 years | 375 | 27 | Boston Vercise RC |
| **P09** | 50-55 | F | 8 | 3 months | 1339 | 25 | Medtronic Activa RC |
| **P10** | 70-75 | F | 7 | 1.8 years | 399 | N/A | Medtronic Percept PC |
| **P11** | 65-70 | M | 9 | 8 months | 1225 | 7 | Boston Vercise Genus |
| **P12** | 65-70 | M | 19 | 1 month | N/A | N/A | Medtronic |

### Dataset

We recorded data from routine deep⍰brain stimulation (DBS) programming sessions at Charing Cross Hospital in London, UK. Individuals with Parkinson’s disease who had subthalamic nucleus (STN)-targeted DBS implants were recruited, provided with study information, and gave written consent. A dry⍰electrode wireless EEG headset (DSI⍰7, Wearable Sensing) was used to record six EEG channels at 300□Hz. EEG was recorded continuously while clinicians adjusted DBS parameters such as contact configuration, stimulation amplitude, distribution, frequency, and pulse width based on clinical judgement and patient response [27].

We included sessions with sufficient parameter variation to generate at least 5,000 training samples, yielding 22 hemispheres across 13 sessions of 12 patients. We analysed data recorded during both arm and hand movements (finger tapping, hand open and close, and hand pronation-supination) and rest periods, for task agnostic classification.

Post-hoc imaging showed active contacts across the STN, adjacent white matter, and in one case the GPi (Figure 1). Active contacts were defined as those tested during the programming session. Both monopolar and bipolar stimulation were present, sometimes within the same session. Each recording was annotated with stimulation parameters (amplitude, contact, frequency, pulse width) for both electrodes.

**Figure 1.**
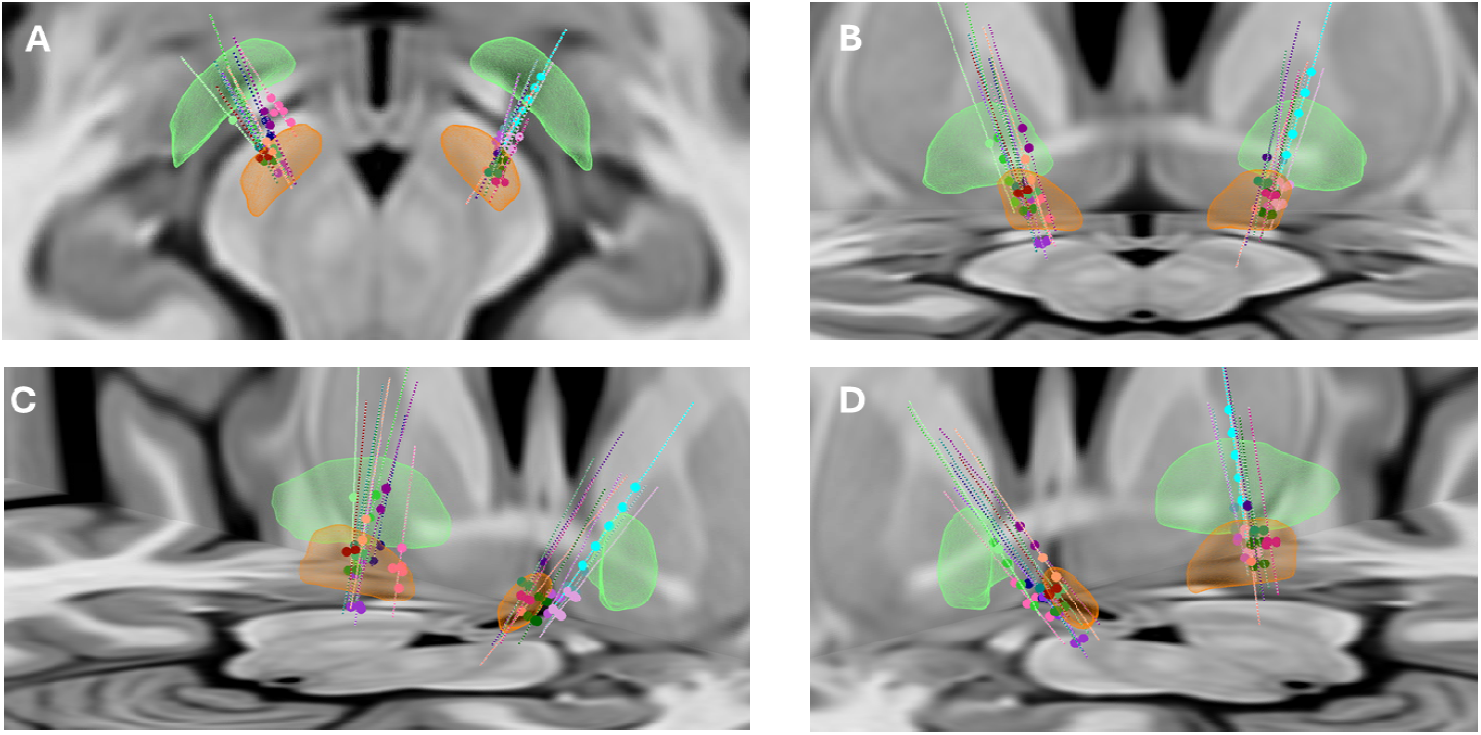
Electrode placement. Visualisation of the DBS electrodes through co-registering of MRI and CT scans by means of advanced normalisation tools with subcortical refine [28,29]. After PaCER [30] algorithm-based localisation, electrode position was manually tuned for a precise estimate. The GPi and STN are represented in green and orange respectively. Each electrode is represented by a unique colour. **A:** Superior plane **B:** Lateral left plane **C:** Lateral right plane **D:** Posterior plane.

### Preprocessing

EEG was bandpass filtered (4–91 Hz, 5th-order Butterworth) and notch filtered at 50 Hz (IIR) to remove electrical noise. Frames without task annotations were discarded. To create “same” and “different” pairs, we implemented a custom Python algorithm with the following conditions: (i) parameters constant within each 1-s segment; (ii) all but one parameter equal between segments; (iii) temporal separation of 2 s–15 min between segments; and (iv) identical task (rest or movement) to avoid confounding task-related activity. Pairs with identical parameters were labeled “same,” while pairs differing in amplitude (≥0.3 mA) or contact were labeled “different.” For contact comparisons, amplitude was not constrained, as its effects vary by contact, and which was validated in the Results. Changes in frequency and pulse width were rare and therefore excluded.

Segments were generated with 50% overlap (o□) and paired with ≤30% overlap (o□) across second segments; no overlap was allowed within pairs. Although this introduced redundancy, it increased sample size and served as data augmentation. The dataset was balanced for class (“same” vs “different”) and for task proportions between classes. Patients were included if they had changes in amplitude or contact, and only the parameters with changes were analysed.

Data was first split into train and test sets, ensuring no overlap between train and test sets. The train set was further divided into train and validation, with a 30% overlap (o_3_) between train and validation sets. To address variable recording lengths, which yielded variable test-set size between patients, we applied k-fold cross-validation, with the number of folds *k* chosen per patient to yield 2,000 test samples each.

### Siamese EEGNet

Models were trained within-patient and independently for each hemisphere and parameter combination that passed the preprocessing steps, yielding 30 independently trained versions of the model. We adapted EEGNet, a compact CNN architecture shown to generalise across diverse EEG tasks [25], into a siamese configuration [31]. Each 1-s EEG segment was processed in parallel using shared weights, producing latent embeddings joined via similarity metrics [32]. The EEGNet architecture performs temporal and spatial convolution separately to extract features from both planes. After the convolutional feature extraction, the embedding of the input is passed through a fully connected layer that creates a vector of dimension *d*_*out*_ = 10, for each of the segments. Then, three distance metrics are calculated and concatenated into a vector of dimension *d*_*sim*_ = 12. The first 10 elements are the element-wise absolute difference between the embedding of the pairs, the 11^th^ element is the dot product of the vectors, and the 12^th^ element is the norm-2 distance between the vectors. The choice of using three distance metrics resulted in a negligible increase in model parameters but led to an increased ability to infer the changes in the data during initial testing of the models. The concatenated vector (of dimension 12×1) is passed through a linear projection layer mapping into a 2-dimensional vector, and finally is passed through a SoftMax and compared to the label. In this embedding, a [0,1] label represents a pair of “same” and a [1,0] label represents a pair of “different” (Figure 2).

**Figure 2.**
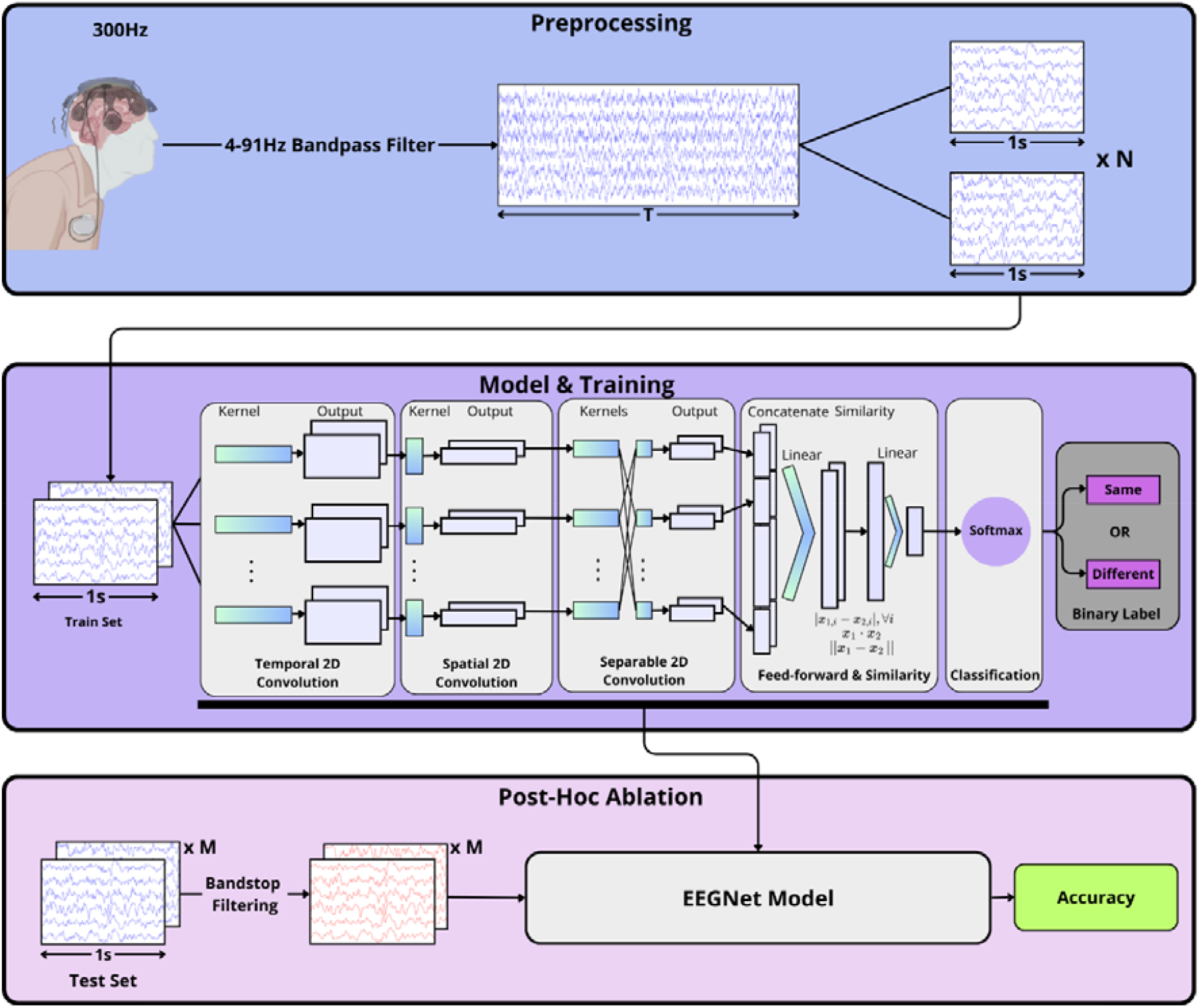
Data processing pipeline. Data processing pipeline divided in three steps. **Preprocessing**: The EEG data is bandpass filtered between 4 to 91Hz and N pairs of 1s EEG segments are generated to train the model. **Model & Training**: Pairs of EEG segments are inputted as an image of 12×300 samples into the model. After convolutional feature extraction, similarity metrics are calculated and a final 2-D embedding that goes through the SoftMax layer to be classified as [0,1] or [1,0] (“same” or “different” respectively). The model dimensions were kept as in the original paper[25]. **Post-Hoc Ablation**: After the model has been trained, features are removed from the test dataset which is passed through the model to compare the accuracy to the baseline with the unfiltered data. When removing a certain feature caused large drops in accuracy, this feature was deemed to be extracted by our model and to be essential for the correct classification of the input. DBS patient schematic created in BioRender.com.

The model was implemented in PyTorch [33]. We used *F*_*l*_ = 8, *D* = 2, which correspond to the number of temporal and spatial filters respectively, and a dropout probability of 0.45 was added. Optimisation used binary cross-entropy loss, Adam [34], and L2 regularisation. Regularisation and learning rate were tuned per patient/parameter combination with a Hyperband scheduler [35] using 30 starting points, implemented in Ray [36]. Models were trained for 450 epochs, with the epoch yielding lowest validation loss selected for testing. To mitigate overfitting, folds with validation accuracy below a variable threshold were excluded, retaining the top 75% unless ≥80% accuracy was achieved. Training was performed within-patient for one parameter at a time (e.g. left electrode amplitude), and performance was evaluated by accuracy.

### Ablation Studies

To identify the frequency bands most critical for distinguishing DBS parameter changes, we performed ablation tests on the trained models [32]. Each band was removed from the test data using 5th-order Butterworth bandstop filters, and model accuracy was re-evaluated. We examined theta–alpha (4–12 Hz), beta (13–30 Hz), low-gamma (31–59 Hz), and mid-gamma (60–90 Hz) bands. To achieve finer spectral localisation, we also applied moving 12-Hz bandstop filters (5th-order Butterworth) sliding from 4 Hz to 91 Hz in 1-Hz steps. Analyses were performed separately for each patient and parameter. A large accuracy drop following removal of a frequency range was interpreted as evidence that the band contained features essential to detecting DBS parameter changes.

### Clustering Analysis

We clustered hemispheres based on the physiological band ablation results, using the drop in model accuracy after band removal (relative to the full-spectrum baseline) as input features. Both K-means and hierarchical clustering were applied, with the optimal *K* determined via scikit-learn’s silhouette score [37]. Clusters were visualised in 2D subspaces of relevant frequency bands to assess separation quality. To examine feature redundancy, we computed Spearman correlations between accuracy drops for the different frequency bands across hemispheres.

### Subharmonic Entrainment

During the initial evaluation of our patients’ PSDs, we identified 5 out of 22 hemispheres which displayed signs of subharmonic entrainment to exactly half of the stimulation frequency. To assess whether this feature was being learned by our models, we separately plotted the moving filter band-stop results for each hemisphere, assessing the relationship between the FTG frequency and the accuracy drop.

## Results

### Classification Results

We trained independent models for the different hemispheres (n=22) and the different types of changes – contact or amplitude. 20 out of 22 hemispheres had a model for amplitude, while only 10 had contact changes, which enabled a contact model. Our models achieved an overall accuracy of 78±9% (mean±SD) in classifying the pairs of EEG data with the binary label “same” or “different”, where “same” refers to both EEG segments in a pair having the same stimulation parameters, and “different” refers to the parameter of interest being different between the two segments. There was no notable difference in accuracy between the changes in amplitude and changes in contact (79±8% and 76±10% for amplitude and contact, respectively, Supplementary Table 2). To ensure the stimulation amplitude doesn’t drive the classification of contact changes, we tested the model’s performance in discriminating pairs of “different” contacts with varying amplitude differences (Supplementary Figure 1). The subset of pairs of “different” contacts that had no amplitude difference between the segments, achieved an accuracy of 75%, equivalent to the accuracy with no control for amplitude. This accuracy represents the performance of models that were trained on all data when evaluated on pairs of “different” contacts that have no changes in amplitude. One model had a low accuracy 57%, with a strong bias towards the label “same”, making the “different” accuracy 31.14% (P05 left, see Supplementary Table 3) and was therefore excluded from this analysis step.

### Frequency Ablation Study &Clustering Analysis

An ablation study was used to find the bands learned by the models. Removing the mid-gamma (60-90Hz) had the strongest effect, causing an average decrease in accuracy of 19% across all hemispheres and parameters, resulting in a low average accuracy of 59%. Mid gamma was the most important frequency band in 22 out of 30 cases, and the Theta-Alpha band was the most important in the remaining 8 hemispheres (Supplementary Table 4).

In a search for a pattern in the ablation results, a clustering analysis of the hemispheres according to their EEG-band band-stop filtering results (Supplementary Table 4), revealed 4 clusters. Each cluster had different dominant bands of ablation accuracy drops (Table 2), and we named them accordingly: Theta-Alpha dominant, Frequency Robust, Mid-gamma dominant, and Slow+Mid-Gamma dominant. Those clusters are linearly separable on both Theta-Alpha/Gamma and Theta-Alpha/Beta 2D planes (Figure 3A+B). While we clustered hemispheres (by averaging across the amplitude and contact models of each hemisphere), this linear separation holds for the individual models (contact/amplitude) separately (Supplementary Figure 2). A correlation analysis revealed that all the ablation accuracy drops were strongly correlated between all bands except Beta (Supplementary Figure 3).

**Table 2.**
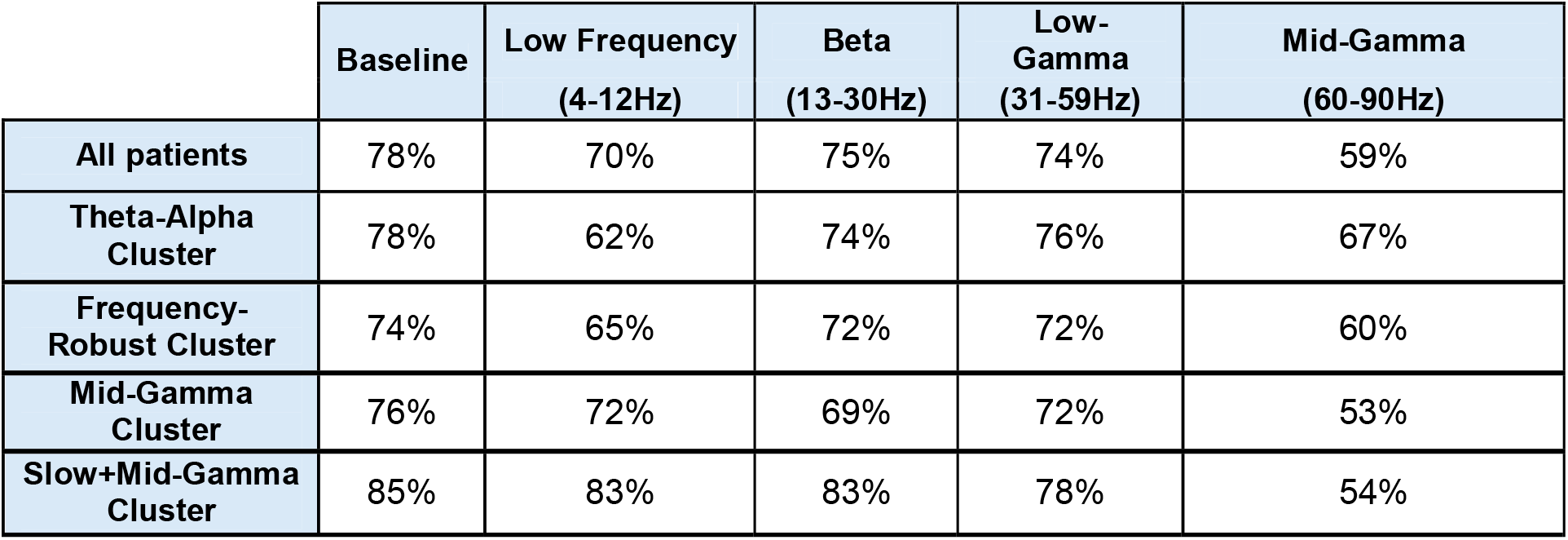
Ablation test accuracies after band-filtering. Accuracy on the test dataset in % for each parameter averaged across hemispheres. The first column represents the baseline accuracy, and each subsequent column represents the ablation accuracies after filtering out one frequency band from the test dataset. For all but one cluster, filtering the mid-gamma (60-90Hz) caused the largest drop in accuracy, making it the most important band for the models’ performance. For a full representation of the accuracy drop per hemisphere and parameter combination, see Supplementary Table 4.

**Figure 3:**
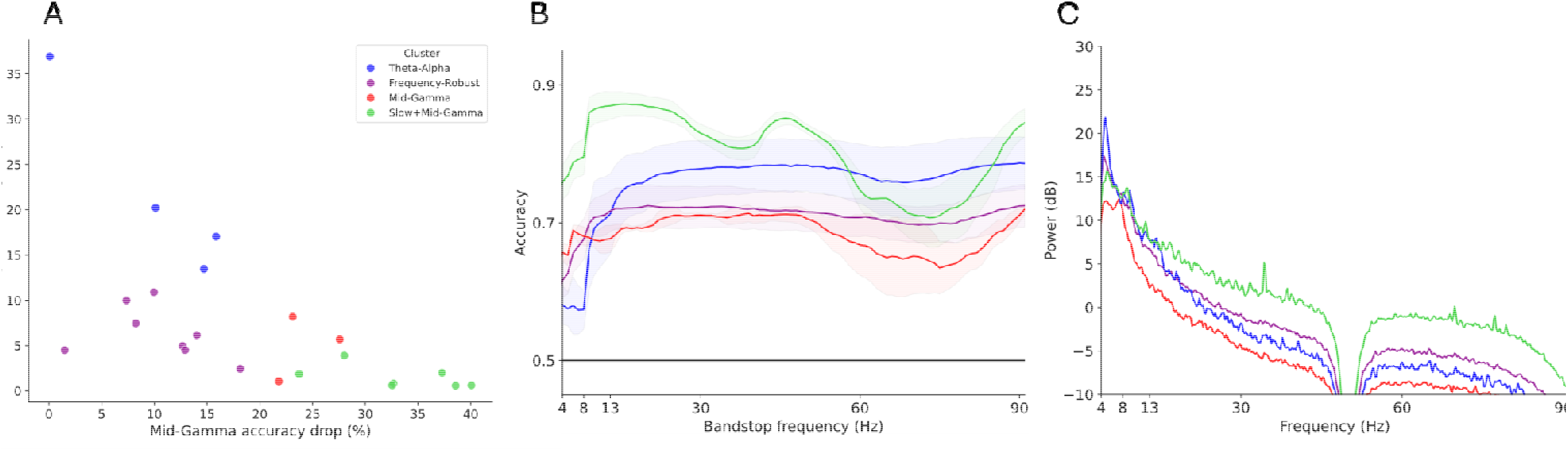
Accuracy results from ablation bandstop study for different patients and parameters. **A:** Each hemisphere plotted in the Mid-Gamma/Theta-Alpha and Beta/Theta-Alpha plane accuracy drop plane colour coded by clusters. Clusters are linearly separable. **B:** A 12Hz bandstop moving filter was swept from 4Hz to 91Hz Accuracy results from the ablation bandstop study of each cluster’s average ± standard error from the mean are plotted in different colours. Legends as in A. **C**: Average Power Spectral Density (PSD) of the input EEG data in dB for each cluster averaged across channels using Welch’s periodogram during the time windows of the pairs of changes in amplitude. Legends as in A.

To further inspect the bands’ contribution to the models, a moving band-stop filtering was used in a secondary ablation analysis. This resulted in a curve tracking the accuracy decay in response to the ablation of different frequencies, which were averaged across models in each cluster (Figure 3C, results for each of the models separately are in Supplementary Figure 4). Here we observe four distinct responses, which are in line with the results from Table 2. Importantly, we observe a drop in the mid-gamma for all clusters, though with varying amplitude. In the Slow+Mid--Gamma group, we observe a drop in the low-gamma range, which is aligned to peaks in the group’s PSD (Figure 3D). In the single hemisphere’s PSDs (Supplementary Figure 4C) of all but one hemispheres in this cluster, we observed a peak in the low-gamma range. Nonetheless, the mid-gamma drop is higher, meaning the most important features being learned for that group lie in the mid-gamma range.

**Figure 4.**
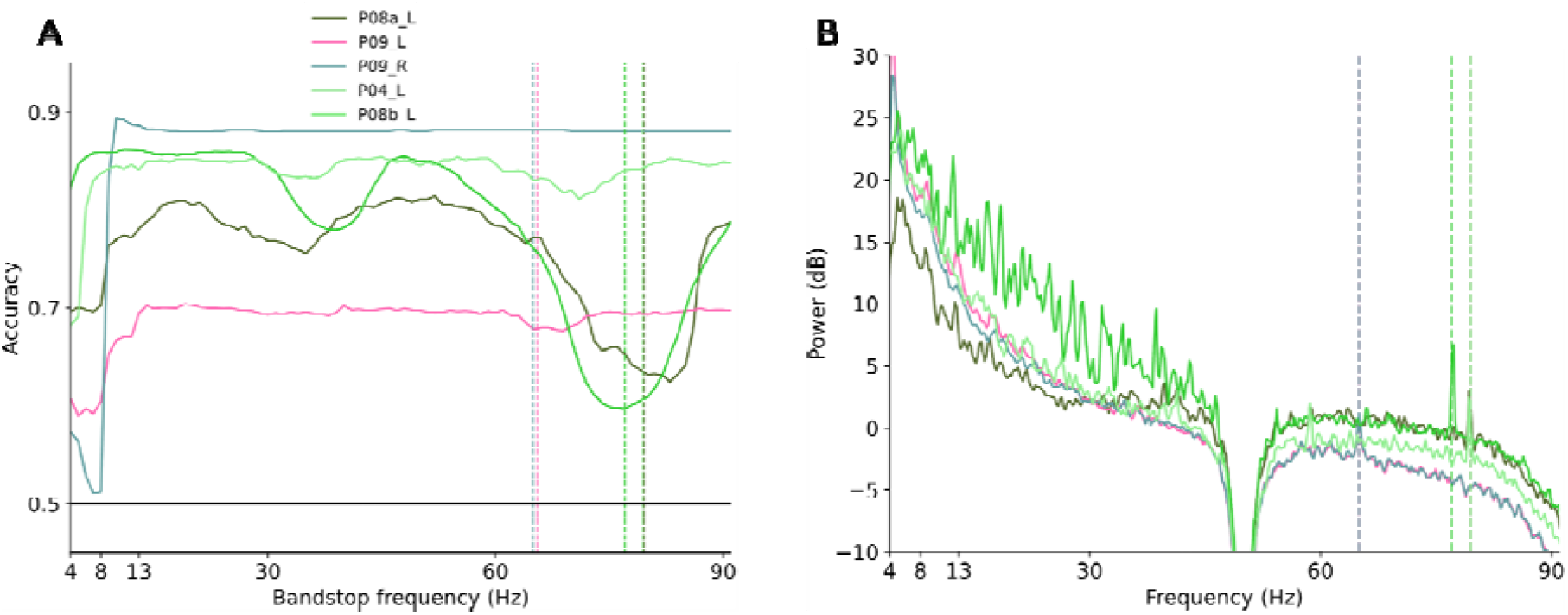
Individual ablation and PSD plots for patients with subharmonic entrainment. **A:** Same protocol as in Figure 3B. Vertical lines represent the subharmonic of the stimulation frequency (i.e.). A small random jitter has been added to the lines to be able to visualise patients that have the same stimulation frequency at the same time. For the specific stimulation frequency values please refer to Supplementary Table 1. **B:** Same protocol as in Figure 3C, with the subharmonic vertical lines added. Colours were chosen to match the group (cluster) each patient belongs to.

### Subharmonic entrainment

The ablation studies highlight the mid-gamma as the band most sensitive to DBS parameter changes, in agreement with many recent studies. However, in most of those it was the FTG and specifically, the subharmonic entrainment that drove it. In our data, only 5 of the 22 hemispheres had subharmonic entrainment (Figure 4). Of those five, only one (P04_L) showed a clear alignment between the drop in accuracy and the stimulation subharmonic (vertical line in Figure 4A). Two other hemispheres (P08a_L and P09_L) showed some alignment, while the remaining two showed no drop in accuracy aligned with the subharmonic of the stimulation frequency (Figure 4A), despite the subharmonic entrainment being present in the PSDs (Figure 4B)

## Discussion

Our siamese CNN on 1-second EEG segments achieved an average accuracy of 78±9% across all 30 models. Using only six-channel dry EEG channels, 1-second windows, and a lightweight deep learning framework, we achieved robust, above-chance performance within 11 out of 12 patients. This represents the first approach capable of discriminating neural responses to small DBS parameter changes at the scalp using a siamese CNN, in a task-agnostic paradigm that mixed rest and movement data. Despite EEG’s lower signal-to-noise ratio, compared to LFPs, its brain-wide coverage enables detection of DBS-related changes, supporting its potential for identifying cortical biomarkers. Those robust performances over very short 1-second windows pave the way towards fast automatic scanning of the vast parameter space of DBS, towards biomarker-guided automated DBS programming [26] and aDBS. While future work will be needed to define the biomarker and correlate it to patient symptoms, our work set the route towards it, with the detection of subtle DBS effects via EEG, which enables automated DBS programming even in patients without LFP-recording-capable devices. Importantly, our work used data from unmodified clinical programming sessions, underscoring its real-world translational relevance.

A key goal of our study was to identify the frequency bands our models extracted to distinguish neural activity at different DBS parameters. Our ablation analyses revealed mid-gamma activity (60–90 Hz) as the most influential frequency band. It produced the largest accuracy drop in 22 of 30 models and a substantial decrease in the remainder, confirming its robustness as a marker of DBS-related neural responses. Removing the entire mid-gamma band (Table 2) caused stronger accuracy losses than those seen in the continuous bandstop plots (Figure 3B), suggesting that specific mid-gamma sub-bands carry distinct information, though the broader range remains important.

Recent DBS studies have started to shift from beta to gamma activity as the band of interest for DBS biomarkers [10,15–21], and our findings are consistent with this trend. Notably, amplitude changes as small as 0.3 mA—often subclinical and undetectable by clinical scales such as the UPDRS—were successfully discriminated here, suggesting that mid-gamma oscillations may be used to define a highly sensitive biomarker. The 1:2 entrainment phenomenon, where neural oscillations occur at half the stimulation frequency, has been proposed as a mechanism underlying mid-gamma activity [10,16,18–21,38]. However, we observed clear subharmonic entrainment only in five hemispheres (Figure 4B), despite adequate stimulation amplitudes (Supplementary Table 1) and inclusion of movement tasks known to enhance this effect [19,20]. Moreover, alignment between subharmonic peaks and accuracy drops was evident only in 3 hemispheres (e.g., P04_L), indicating that cortical mid-gamma involvement extends beyond 1:2 entrainment mechanisms.

Theta–alpha oscillations (4–12 Hz) have also been implicated in PD and DBS research as potential markers of motor function, including bradykinesia, tremor, and rigidity [39–44]. Low-frequency changes during DBS ON states have also been reported [41,45–47], though others have found these oscillations unsuitable for describing STN coupling, favouring high-beta instead [48]. Our findings reconcile both perspectives: a subset of hemispheres exhibited strong low-frequency modulation, while others showed minimal effects. Importantly, even within the Theta–Alpha group, removing mid-gamma activity still reduced accuracy (Table 2, Figure 3B), suggesting an interaction between mid-gamma and theta–alpha oscillations in representing DBS-related cortical dynamics. In a recent study using the same dataset, we looked specifically at bursting and reported amplitude-dependent changes in both alpha (8–12 Hz) and mid-gamma (60–90 Hz) bursting [27]. Here, our models were naïve to any bursting definition (requiring decisions on envelopes and thresholds) and in a purely data-driven approach, classified both amplitude and contact changes, indicating that they may have captured bursting features or related spectral dynamics across these frequency ranges.

Despite beta power being a known marker in PD and its modulation by DBS, our results did not consistently support its significance in terms of accuracy impact (Table 2). We also found that the beta range was the only band whose drop in accuracy when filtered did not correlate with any of the other frequency bands. Overall, our findings indicate that mid-gamma oscillations (60–90 Hz) are the dominant cortical signature of DBS parameter changes across patients, with theta–alpha activity (4–12 Hz) providing complementary information in a subset of hemispheres.

Despite demonstrating a proof-of-concept for parameter discrimination at the cortical level in a clinically relevant setting, our work has several limitations. First and foremost, our study focused purely on the sensitivity of the cortical activity to changes in the DBS settings, often changes that are too small to induce a detectable change in symptoms, and thus did not map those to DBS efficacy and symptom changes, which should be assessed in future studies translating our approach. In addition, a key concern was whether the models might misclassify stimulation artefacts as neural responses, given that they were trained without predefined feature constraints. Since changes in the EEG signal as a result of stimulation artefacts are more likely for changes in the stimulation amplitude (or any other feature of the duty cycle) then for changes in the contact, the fact our models achieved 75% accuracy in distinguishing “different” contacts when the amplitude (and all other feature of the duty cycle) remained unchanged is very reassuring (Supplementary Figure 1). Since contacts are only ~0.5 mm apart in deep brain regions, such differences are unlikely to produce measurable EEG artefacts, supporting a neural basis for classification. We also found no bias in classifying monopolar vs. bipolar stimulation, suggesting that our models accurately identify contact points, not stimulation types (Supplementary Tables 5 and 6). Considering the common view that bipolar settings do not affect the cortex with stimulation artefacts [50,51], our results, which show similar accuracies in bipolar pairs, represent true changes in cortical activity and not stimulation artefacts (Supplementary Tables 5 and 6). Another limitation was the restriction to rest and arm movement tasks, which limits the generalizability to daily life and free behaviour. This was done to reduce potential movement artefacts, as tasks like walking introduced strong EEG artefacts. Yet, our models’ success across two different tasks suggests early evidence of task robustness. Finally, as a proof-of-concept study, we used limited explainability methods, focusing on ablation studies. While we found mid-gamma (60-90Hz) critical for model performance across all patients and theta-alpha (4-12Hz) for a subset of hemispheres, with our current methods, we could not pinpoint the exact features extracted and define a biomarker. Future work will focus on improving the AI architecture into an inherently more explainable one to better understand the brain’s neural response, and on mapping it to the DBS effect on motor symptoms.

## Conclusion

In this work, we introduced a novel method for detecting changes in DBS parameters at the cortical level, achieving significantly above chance-level performance. By testing this on 30 independent models on different patients, this work represents a robust proof of concept that we can reliably detect changes in contact and changes in amplitude as small as 0.3mA at the cortical level with only 1s segments of 6 EEG channels. To gain preliminary insight into the neural response to changes in DBS parameters, we performed ablation studies and found that our models were mostly extracting features from the mid-gamma (60-90Hz) to accurately classify pairs of EEG segments as “same” or “different” depending on the DBS parameters each segment was recorded with. However, we found that the 1:2 entrainment phenomenon, which has been emerging in the field as a potential biomarker, was not an important feature for our dataset and models. Despite mid-gamma being the main frequency range where information lies, we saw heterogeneity in the frequency characteristics of the cortical responses across our cohorts. This revealed the theta-alpha oscillations (4-12Hz) as a frequency range where complementary information to the mid-gamma could lie. Overall, we achieved consistent and robust results across our analysis, which involved models trained independently for each hemisphere of each patient. Future work will focus on improving the AI methods, both in terms of accuracy and explainability, and in creating a patient-agnostic model that could be evaluated on a larger sample size with minimal training. Other steps required to bring this work to the clinic will involve using the found biomarkers to predict specific DBS parameters and their effect on symptom severity. Overall, this work sets a new path towards EEG-based biomarker-guided DBS programming for more effective and personalised treatments.

## Data Availability

The dataset used in this project contains sensitive patient data and will therefore not be made publicly available. The authors are keen on collaborative work with this dataset, please contact the corresponding author if interested.

## Notes

### Competing Interest Statement

The authors have declared no competing interest.

### Funding Statement

The study was supported by the Edmond and Lily Safra Fellowship awarded to S.H. and by the UK Dementia Research Institute Care Research & Technology Centre. N.C.P and C.G. are supported by UK Research and Innovation [UKRI Centre for Doctoral Training in AI for Healthcare grant number EP/S023283/1]

### Author Declarations

London - Harrow Research Ethics Committee gave ethical approval for this work

### Summary of Updates

We increased the number of patients and, as a result, trained models, demonstrating the robustness of the approach. With those increased numbers, we were able to draw some conclusions and insights on frequencies of interest. We also rewrote and reframed the manuscript to better deliver its message.

