## Supplementary material for "EEG-based Deep Learning Reveals Cortical Sensitivity to Small Changes in Deep Brain Stimulation Parameters"

### Supplementary Information

| | Left Amplitude (mA) | # of Left Contact Combinations | Left Frequency (Hz) | Left Pulse-Width (µs) | Right Amplitude (mA) | # of Right Contact Combinations | Right Frequency (Hz) | Right Pulse-Width (µs) |
|---|---|---|---|---|---|---|---|---|
| **P01** | 6.3 – 6.8 | 3 | 159 | 80 | 5.3 – 5.8 | 1 | 159 | 70, 80 |
| **P02** | 3.1 – 7.0 | 2 | 179 | 70, 90 | 1.2 – 7.0 | 8 | 179 | 60, 90 |
| **P03** | 0.5 – 2.3 | 2 | 130 | 60 | 0.5 – 3.0 | 3 | 130 | 60 |
| **P04** | 0.5 – 5.3 | 6 | 154 | 60 | 4.4 | 1 | 116 | 50 |
| **P05** | 4.9 | 2 | 145 | 60 | 5.1 – 5.5 | 1 | 145 | 60 |
| **P06** | 4.9 – 5.2 | 1 | 174 | 60 | 3.5 – 3.8 | 1 | 174 | 60 |
| **P07** | 3.0 – 9.5 | 2 | 125 | 30, 40, 60 | 1.5 – 3.4 | 1 | 125 | 40, 60 |
| **P08a** | 3.9 – 5.9 | 2 | 159 | 60 | 0.7 – 1.7 | 1 | 119 | 50 |
| **P08b** | 5.8 – 6.9 | 1 | 159 | 60, 70 | 1.7 | 1 | 119 | 50 |
| **P09** | 1.0 – 4.0 | 5 | 130 | 60 | 0.6 – 2.5 | 2 | 130 | 60 |
| **P10** | 3.0 – 3.5 | 1 | 130 | 60 | 1.2 – 2.2 | 1 | 130 | 60 |
| **P11** | 2.8 – 3.8 | 1 | 130 | 60 | 3.2 – 4.2 | 1 | 130 | 50 |
| **P12** | 0.3 – 1.1 | 1 | 125 | 60 | 0.3 – 1.5 | 2 | 125 | 50, 60 |

***Supplementary Table 1. Parameters range across the different patients.*** *Parameters range for each parameter and patient. The number in the contact cells represents the number of unique contact configurations that were displayed, and in the amplitude cell, the range is represented. However, the changes in the amplitude were not linear or systematic, meaning that a big range does not necessarily result in more unique amplitude configurations.*

| | Amplitude (%) | Contact (%) | Average (%) |
|---|---|---|---|
| **P01 Left** | 77.85 | 77.20 | 77.53 |
| **P01 Right** | 80.19 | N/A | 80.19 |
| **P02 Left** | 72.75 | 90.16 | 81.46 |
| **P02 Right** | 70.07 | 73.11 | 71.59 |
| **P03 Left** | 76.23 | 75.62 | 75.93 |
| **P03 Right** | 79.05 | 67.88 | 73.47 |
| **P04 Left** | 84.89 | 88.33 | 86.61 |
| **P05 Left** | N/A | 57.57 | 57.57 |
| **P06 Left** | 80.31 | N/A | 80.31 |
| **P06 Right** | 74.17 | N/A | 74.17 |
| **P07 Left** | 69.80 | N/A | 69.80 |
| **P07 Right** | 80.23 | N/A | 80.23 |
| **P08a Left** | 80.23 | N/A | 80.23 |
| **P08b Left** | 85.20 | N/A | 85.20 |
| **P09 Left** | 69.16 | 72.04 | 70.6 |
| **P09 Right** | N/A | 88.07 | 88.07 |
| **P10 Left** | 86.85 | N/A | 86.85 |
| **P10 Right** | 94.80 | N/A | 94.80 |
| **P11 Left** | 89.86 | N/A | 89.86 |
| **P11 Right** | 88.94 | N/A | 88.94 |
| **P12 Left** | 71.59 | N/A | 71.59 |
| **P12 Right** | 59.2 | 72.47 | 65.84 |
| **Average** | 78.58 | 76.25 | **77.80** |

***Supplementary Table 2: Test accuracies across patients and parameters.*** *Classification accuracy for changes in contact and amplitude of both hemispheres. Across every patient and parameter, we achieved an accuracy of 77.80%, showing a proof-of-concept that the model can accurately distinguish between stimulation amplitude and contact points at the cortical level, with amplitude changes as small as 0.3mA.*

| | Same (%) | Different (%) | Average (%) |
|---|---|---|---|
| **P01 Left** | 66.67% | 88.15% | 77.53 |
| **P01 Right** | 67.94% | 89.86% | 80.19 |
| **P02 Left** | 74.95% | 93.14% | 81.46 |
| **P02 Right** | 76.73% | 66.35% | 71.59 |
| **P03 Left** | 72.14% | 81.00% | 75.93 |
| **P03 Right** | 62.84% | 71.98% | 73.47 |
| **P04 Left** | 85.11% | 88.19% | 86.61 |
| **P05 Left** | 31.14% | 84.00% | 57.57 |
| **P06 Left** | 74.78% | 85.84% | 80.31 |
| **P06 Right** | 66.37% | 81.98% | 74.17 |
| **P07 Left** | 68.63% | 70.98% | 69.80 |
| **P07 Right** | 77.27% | 83.19% | 80.23 |
| **P08a Left** | 72.05% | 88.72 % | 80.23 |
| **P08b Left** | 79.22% | 91.18% | 85.20 |
| **P09 Left** | 69.92% | 70.76% | 70.6 |
| **P09 Right** | 77.29% | 98.86% | 88.07 |
| **P10 Left** | 81.19% | 92.53% | 86.85 |
| **P10 Right** | 93.44% | 94.06% | 94.80 |
| **P11 Left** | 84.43% | 95.29% | 89.86 |
| **P11 Right** | 81.34% | 96.54% | 88.94 |
| **P12 Left** | 94.55% | 48.64% | 71.59 |
| **P12 Right** | 83.99% | 47.82% | 65.84 |
| **Average** | 74.64 | 82.23 | **77.80** |

***Supplementary Table 3: Test accuracies across hemispheres by label.*** *Classification accuracy by class (“same” or “different”) averaged across contact and amplitude. Shown per hemisphere.*

| | Baseline | Mid-GammaGamma (60-90Hz) | Low Gamma (31-59Hz) | Beta (13-30Hz) | Theta-Alpha (4-12Hz) |
|---|---|---|---|---|---|
| P01 Left | 77.85 | 47.55 | 74.05 | 69.63 | 71.94 |
| P01 Left | 80.18 | 62.04 | 78.51 | 79.12 | 77.74 |
| P01 Right | 77.2 | 52.33 | 71.73 | 73.07 | 71.73 |
| P02 Left | 72.75 | 65.62 | 73.34 | 72.66 | 67.68 |
| P02 Left | 70.07 | 59.08 | 70.92 | 67.76 | 53.42 |
| P02 Right | 90.16 | 71.96 | 88.78 | 90.05 | 85.29 |
| P02 Right | 73.11 | 54.69 | 67.65 | 64.08 | 62.82 |
| P03 Left | 76.23 | 56.07 | 57.21 | 57.14 | 53.07 |
| P03 Left | 79.05 | 65.88 | 75.88 | 74.82 | 57.11 |
| P03 Right | 75.62 | 59.71 | 79.36 | 77.14 | 74.54 |
| P03 Right | 67.88 | 65.74 | 73.53 | 68.29 | 54.32 |
| P04 Left | 84.89 | 59.18 | 66.93 | 65.72 | 60.07 |
| P04 Left | 88.33 | 50.82 | 77.01 | 83.75 | 84.62 |
| P05 Left | 57.57 | 57.03 | 80.1 | 88.65 | 86.98 |
| P06 Left | 80.31 | 64.45 | 79.28 | 71.61 | 63.27 |
| P06 Right | 74.17 | 51.05 | 69.97 | 66.74 | 65.99 |
| P07 Left | 69.8 | 61.57 | 69.8 | 67.25 | 62.35 |
| P07 Right | 80.23 | 72.89 | 78.75 | 79.18 | 70.24 |
| P08a Left | 80.38 | 52.37 | 75.32 | 78.40 | 76.47 |
| P08b Left | 85.2 | 61.47 | 82.45 | 84.22 | 83.33 |
| P09 Left | 69.16 | 58.57 | 67.34 | 69.94 | 59.03 |
| P09 Left | 72.04 | 62.69 | 69.35 | 70.46 | 60.37 |
| P09 Right | 88.07 | 88.15 | 88.07 | 88.07 | 51.14 |
| P10 Left | 86.85 | 49.57 | 79.33 | 85.71 | 84.85 |
| P10 Right | 94.8 | 62.24 | 70.2 | 90.56 | 94.18 |
| P11 Left | 89.86 | 49.8 | 85.55 | 87.81 | 90.47 |
| P11 Right | 88.94 | 50.37 | 83.91 | 87.89 | 89.52 |
| P12 Left | 71.59 | 49.79 | 65.72 | 61.79 | 70.55 |
| P12 Right | 59.2 | 52.81 | 57.32 | 57.05 | 55.01 |
| P12 Right | 72.47 | 53.02 | 71.5 | 69.29 | 67.62 |
| Average | 77.80 | 58.95 | 74.30 | 74.93 | 70.19 |

***Supplementary Table 4. Full band-filtering results.*** *Accuracy in % after filtering different bands from the test set with a model trained on the full data. The results are shown for every patient and parameter that was analysed in this study. For 22 out of 30 cases, the mid-gamma (60-90Hz) caused the most significant drop in accuracy. Forthe remaining hemispheres, the Low Frequency band (4-12Hz) caused the highest drop in accuracy, but the mid-gamma still caused a significant decrease.*

| | | Same M-M | Same B-B | Diff M-M | Diff M-B | Diff B-B |
|---|---|---|---|---|---|---|
| **1** | P01 Left | N/A | 88.00% N=750 | N/A | N/A | 66.40% N=750 |
| **2** | P02 Left | N/A | 96.72% N=945 | N/A | N/A | 83.60% N=945 |
| **3** | P02 Right | 71.24% N=445 | 87.73% N=269 | 65.24% N=607 | 90.00% N=100 | 71.43% N=7 |
| **4** | P03 Left | 77.16% N=648 | N/A | 74.07% N=648 | N/A | N/A |
| **5** | P03 Right | 74.07% N=540 | N/A | 62.22% N=540 | N/A | N/A |
| **6** | P04 Left | 8.82% N=34 | 88.44% N=926 | 94.25% N=87 | 91.04% N=603 | 90.00% N=270 |
| **7** | P05 Left | 84.00% N=700 | N/A | 31.14% N=700 | N/A | N/A |
| **8** | P09 Left | 67.22% N=540 | N/A | 76.85% N=540 | N/A | N/A |
| **9** | P09 Right | 98.86% N=612 | N/A | 77.29% N=612 | N/A | N/A |
| **10** | P12 Right | 65.30% N=928 | N/A | 79.63% N=928 | N/A | N/A |

***Supplementary Table 5: Accuracy and number for each patient's hemisphere separated by polarity in pairs of "same" or "different" contact.*** *M-M means that the patients received monopolar stimulation in both segments of the pair. M-B means one segment had monopolar stimulation and the other bipolar stimulation. B-B means the patient received bipolar stimulation in both segments. This shows that our models can learn to differentiate contact even when the stimulation type is the same across both segments of a pair, and that our models can learn regardless of the stimulation type. Additionally, it shows that our data mostly had same stimulation type pairs and that our models were able to learn when presented with both stimulation types at the same time.*

| | Same M-M | Same B-B | Diff M-M | Diff B-B |
|---|---|---|---|---|
| P01 Left | N/A | 87.64% N=736 | N/A | 68.07% N=736 |
| P01 Right | N/A | 92.38% N=656 | N/A | 67.99% N=656 |
| P02 Left | N/A | 86.52% N=512 | N/A | 58.98% N=512 |
| P02 Right | 46.19% N=578 | 86.81% N=182 | 84.21% N=760 | N/A |
| P03 Left | 82.11% N=570 | N/A | 70.35% N=570 | N/A |
| P03 Right | 85.56% N=540 | N/A | 73.52% N=540 | N/A |
| P04 Left | 100.00% N=21 | 90.66% N=899 | 61.54% N=26 | 79.42% N=894 |
| P06 Left | 85.84% N=678 | N/A | 74.78% N=678 | N/A |
| P06 Right | 81.98% N=666 | N/A | 66.37% N=666 | N/A |
| P07 Left | 24.26% N=169 | 84.23% N=596 | 76.64% N=458 | 56.68% N=307 |
| P07 Right | 83.19% N=946 | N/A | 77.27% N=946 | N/A |
| P08a Left | 88.72% N=780 | N/A | 72.05% N=780 | N/A |
| P08b Left | 91.18% N=510 | N/A | 79.22% N=510 | N/A |
| P09 Left | 73.25% N=770 | N/A | 65.06% N=770 | N/A |
| P10 Left | 92.42% N=924 | N/A | 81.28% N=924 | N/A |
| P10 Right | 94.39% N=980 | N/A | 95.20% N=980 | N/A |
| P11 Left | 95.29% N=488 | N/A | 84.43% N=488 | N/A |
| P11 Right | 96.54% N=954 | N/A | 81.34% N=954 | N/A |
| P12 Left | 48.64% N=954 | N/A | 94.55% N=954 | N/A |
| P12 Right | 29.96% N=908 | N/A | 88.44% N=908 | N/A |

***Supplementary Table 6: Accuracy and number for each patient's hemisphere separated by polarity in pairs of "same" or "different" amplitude****. M-M means that the patients received monopolar stimulation in both segments of the pair. B-B means the patient received bipolar stimulation in both segments. Since the contact was the same in two segments of a pair of amplitude change, the M-B case is not present. This shows that are models can learn to differentiate amplitude regardless of the stimulation type. Additionally, it shows that our models were able to learn when presented with both stimulation types at the same time.*

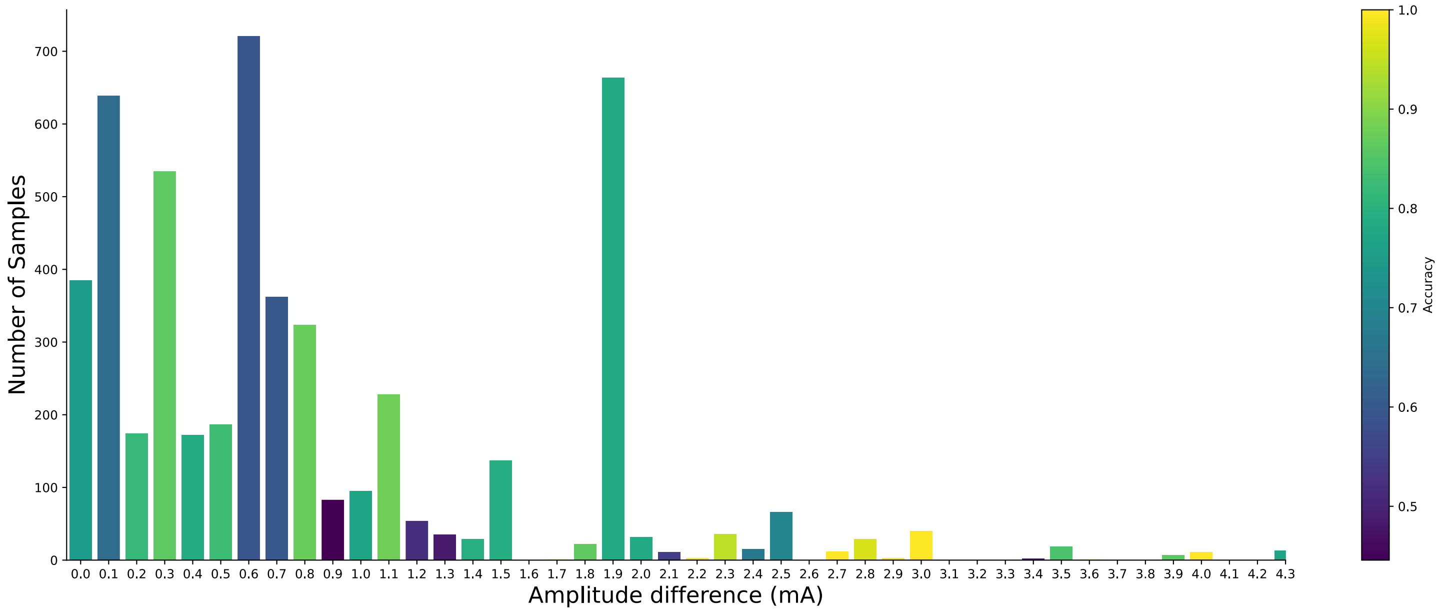

***Supplementary Figure 1: Accuracy distribution in pairs of "different" contact with different amplitude changes***. *The different sample sizes are represented as bars, where the colour represents the accuracy varying from 0 to 1. Our model achieves 75.06% accuracy when no changes in amplitude are present with a sample size of N = 385 pairs. We can see that our models are able to differentiate changes in contact across a wide range of amplitude differences. Only P01-P05 had changes in contact with no changes in amplitude. P05 was excluded from this study as it was the patient with almost chance-level accuracy (57.5%) and when inspecting the confusion matrix we found that this model had a strong bias towards the label "same", making the accuracy for pairs of "different" 31.14%.*

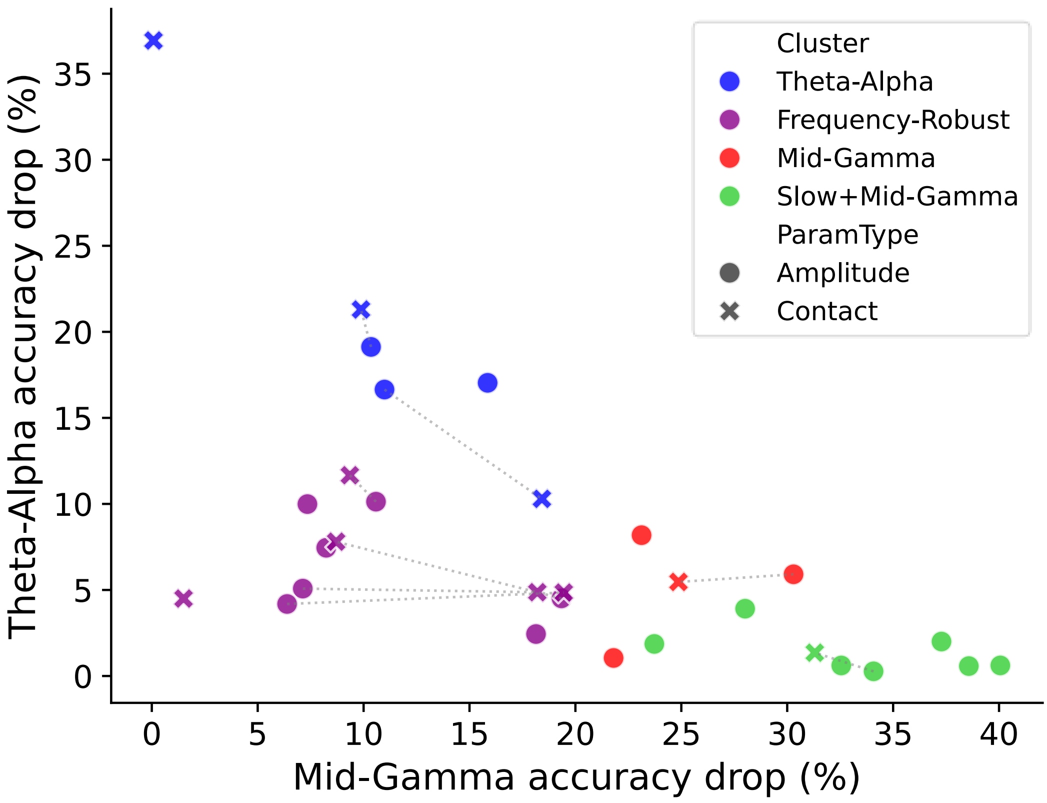

***Supplementary Figure 2: Clustering of different hemispheres/parameter combinations in different frequency drop planes.*** *Same protocol as in Figure 3A. Here we plotted each parameter and hemisphere combination, with the colour corresponding to the clusters given using averages across parameters. Amplitude and contact are represented by circles and crosses respectively. We see that the clustering properties are conserved after plotting each parameter separately, suggesting small differences in the frequency characteristics of the cortical response between changes in amplitude and contact of the same hemisphere.*

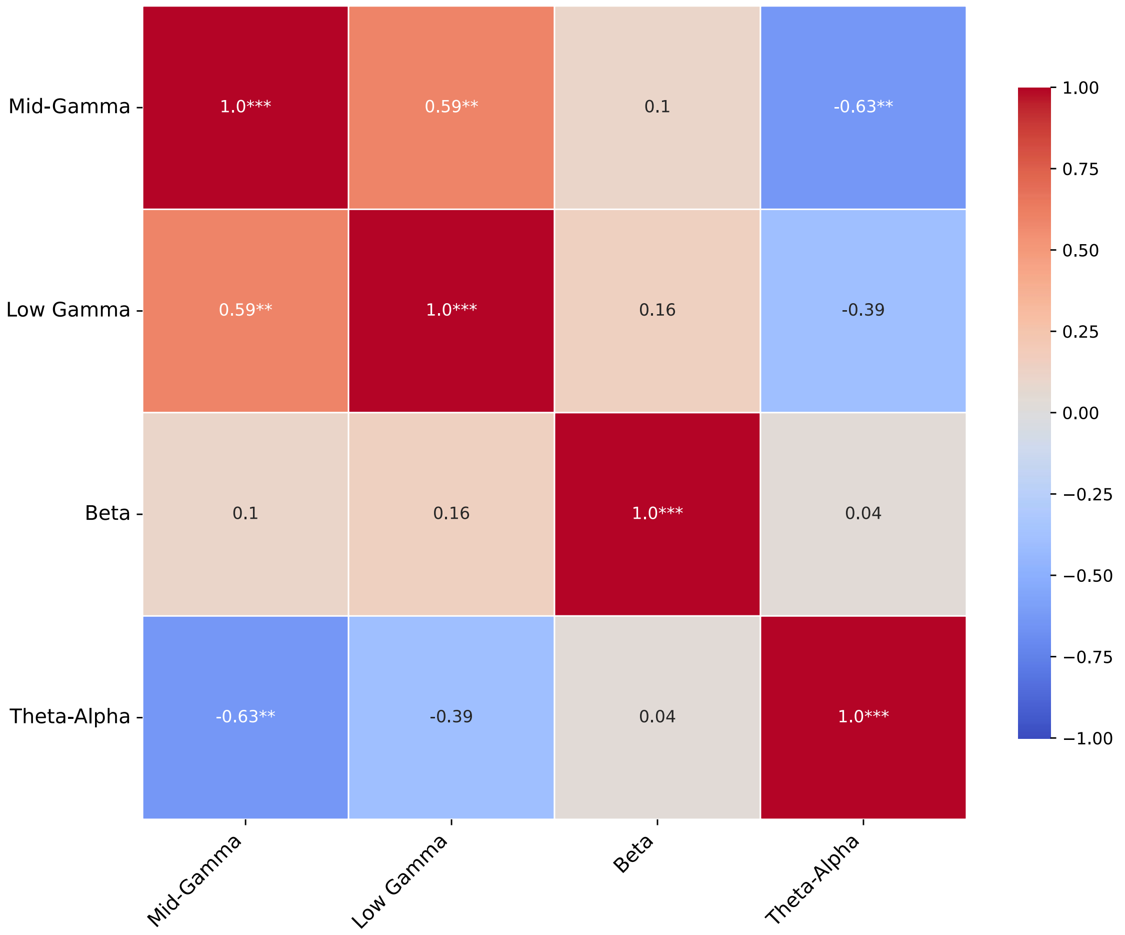

***Supplementary Figure 3: Correlation matrix of the accuracy drop after filtering out different frequency bands across hemispheres.*** *The correlation coefficient is reported and visually represented by the colour as per the colour bar. Statistical significance is represented with *: p<0.05, **:p<0.01, ***p<0.001.*

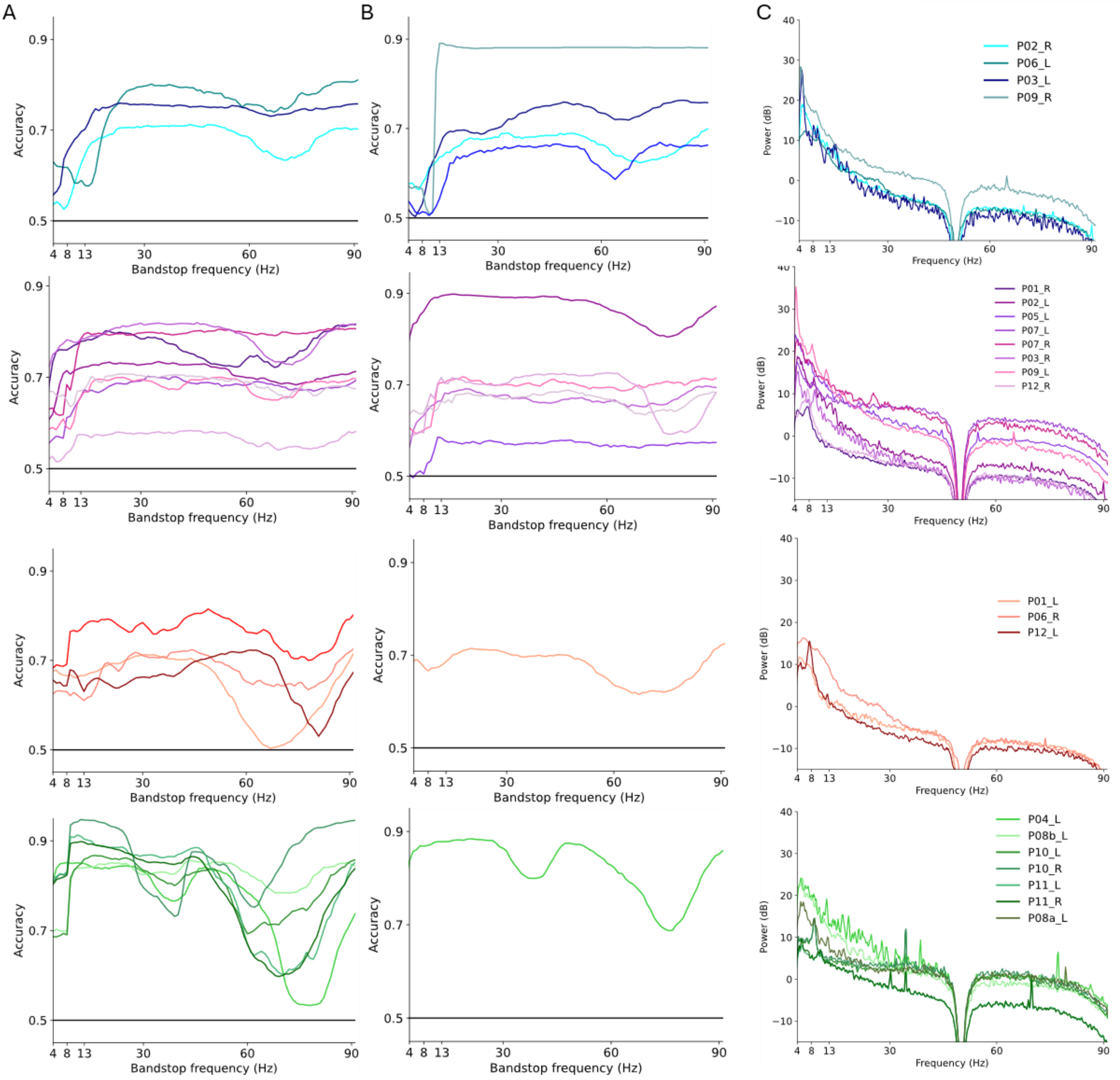

***Supplementary Figure 4. Individual Ablation results for each hemisphere and parameter change, grouped by clusters.*** **A,B:** *Accuracy results with a 12Hz bandstop moving filter, as in Figure 3B, but with each hemisphere plotted as an individual line for changes in Amplitude (**A**) and Contact (**B**).* **C:** *Average Power Spectral Density (PSD), as in Figure 3C, but with each individual line representing a hemisphere. The rows represent the Theta-Alpha, Frequency-Robust, Mid-Gamma and Slow+Mid-Gamma clusters, respectively.*
